# Juvenile Mucopolysaccharidosis plus disease caused by a missense mutation in *VPS33A*

**DOI:** 10.1101/2022.08.27.22279208

**Authors:** Elena V. Pavlova, Dorit Lev, Marina Michelson, Keren Yosovich, Hila Gur Michaeli, Nicholas A. Bright, Paul T. Manna, Veronica Kane Dickson, Karen L. Tylee, Heather J. Church, J. Paul Luzio, Timothy M. Cox

## Abstract

**Background:** A rare and fatal disease resembling mucopolysaccharidosis in infants, is caused by impaired intracellular endocytic trafficking due to deficiency of core components of the intracellular membrane-tethering protein complexes, HOPS and CORVET.

**Methods:** Whole Exome Sequencing identified a novel VPS33A mutation in a patient suffering from a variant form of mucopolysaccharidosis. Electron and confocal microscopy, immunoblotting, and glycosphingolipid trafficking experiments were undertaken to investigate the effects of the mutant VPS33A in patient-derived skin fibroblasts.

**Results:** We describe an attenuated juvenile form of VPS33A-related syndrome - mucopolysaccharidosis plus in a man who is homozygous for a hitherto unknown missense mutation (NM_022916.4: c.599 G>C; R200P) in a conserved region of the VPS33A gene. Urinary glycosaminoglycan analysis revealed increased heparan, dermatan sulphates and hyaluronic acid. We showed decreased abundance of VPS33A in patient derived fibroblasts and provided evidence that the R200P mutation leads to destabilisation of the protein and proteasomal degradation. As in the infantile form of mucopolysaccharidosis plus, the endocytic compartment in the fibroblasts also expanded – a phenomenon accompanied by increased endolysosomal acidification and impaired intracellular glycosphingolipid trafficking. Experimental treatment of the patient’s cultured fibroblasts with the proteasome inhibitor, bortezomib, or exposure to an inhibitor of glucosylceramide synthesis, eliglustat, improved glycosphingolipid trafficking.

**Conclusion:** To our knowledge this is the first report of an attenuated juvenile form of VPS33A insufficiency characterised by appreciable residual endosomal-lysosomal trafficking and a milder mucopolysaccharidosis plus than the disease in infants. Our findings expand the proof of concept of redeploying clinically approved drugs for therapeutic exploitation in patients with juvenile as well as infantile forms of mucopolysaccharidosis plus disease.

## INTRODUCTION

A fatal lysosomal disease resembling mucopolysaccharidosis, caused by a missense mutation in VPS33A gene NM_022916.5:c.1492C>T (c.1492C>T); NP_075067.2:p.Arg498Trp (p.R498W), has recently been identified in infants of Turkish and Yakutian ethnic background: this has been named mucopolysaccharidosis plus (MPSPS, MIM: 610034) (Dursun,2017; Kondo, 2017; Pavlova, 2019). The mutation was found in homozygous form in two infant siblings born to a consanguineous Turkish family (Dursun, 2017) and in 17 young children in the Republic of Sakha Yakutia, an ethnically Turkic population originating from nomadic people who migrated across the Balkans and Central Asia (Kondo, 2017; Pavlova, 2019; Vasilev, 2020). The disease is apparent in early infancy with manifestations suggesting a mucopolysaccharidosis: coarse features, multiple dysostosis, breathing difficulties, joint contractures, impaired motor development, diffuse muscle hypotonia, ataxia and inability to stand or walk. Of note, however, systemic manifestations shared with the Chédiak-Higashi, Hermansky-Pudlak and Griscelli syndromes that affect lysosome-related organelles are found: these include broncho-pulmonary complications, severe renal impairment, susceptibility to infections, defective immunity associated with neutropenia and appearance of azurophilic granules in peripheral lymphocytes, hypopigmentation of retina, nystagmus and a bleeding tendency. Hitherto, all Yakutian patients reported with this disease in infancy have died by the age of 4 years (Kondo, 2017; Pavlova, 2019; Vasilev, 2020), the Turkish patients died before they were 6 years old (Dursun,2017). In consideration of the characteristic clinical features of mucopolysaccharidosis, biochemical investigations revealed pathological excess of urinary glycosaminoglycans and sialylated conjugates while activities of the cognate lysosomal hydrolases responsible for degradation of these complex mucopolysaccharides were unimpaired. In culture, fibroblasts from the patients showed increased vacuolation and expansion of the lysosomal compartment. Defective endocytic trafficking of glycosphingolipids was also identified (Pavlova, 2019), - a feature typical of sphingolipidoses regardless of their genetic cause (Pagano, 2003; Puri, 1999; te Vruchte, 2004; Pryor, 2006; Marks, 2013).

The disease is caused by a reduced abundance of vacuolar protein sorting 33A protein (VPS33A), a component of two intracellular, hexameric, membrane-tethering, protein complexes, the homotypic fusion and vacuole protein sorting complex (HOPS) and the class C core vacuole/endosome tethering complex (CORVET). These complexes regulate SNARE assembly and tether endosomes and lysosomes in the endocytic pathway (Brocker, 2012; Seals, 2000; Perini, 2014; Balderhaar, 2013). Genetic deficiency of the HOPS/CORVET subunit VPS16 has also been shown to phenocopy mucopolysaccharidosis plus disease (Sofou, 2021). We showed that treatment with a proteasome inhibitor rescued the mutant VPS33A^R498W^ from proteasomal degradation in experimental cell lines expressing the protein variant (Pavlova, 2019). We explored the effects of two clinically approved drugs on the abnormal late endocytic trafficking in patient-derived fibroblasts: (i) the proteasome inhibitor, bortezomib, to counter the premature destruction of unstable mutant VPS33A molecules and (ii) inhibition of glucosylceramide biosynthesis with the UDP-glucose: N-acylsphingosine D-glucosyltransferase (UGCG) inhibitor, eliglustat tartrate (Pavlova, 2019). Each treatment substantially improved defective intracellular trafficking of glycosphingolipids and thus offer a potential therapeutic opportunity in this lethal orphan disease (Pavlova, 2019) [3]. Moreover, should substrate reduction with eliglustat (Shayman, 2015; Cox, 2010; McEachern, 2007) ameliorate the systemic disease, an opportune case for using brain-penetrant analogues (Marshall, 2016; Fujii, 2021) to address the neurodegenerative manifestations of the MPS plus would be greatly strengthened.

For the first time here we describe a unique VPS33A-related cellular trafficking syndrome - mucopolysaccharidosis plus in an adult who is homozygous for a hitherto unknown missense mutation (NM_022916.4: c.599 G>C; R200P) in a conserved region of the VPS33A gene. Based on the age of presentation, longevity, absence of severe systemic complications and susceptibility to infections, biochemical and cellular pathology in the context of the variant, we contend that the mutation is associated with an attenuated juvenile form of the mucopolysaccharidosis plus disease.

## MATERIALS AND METHODS

### Patients

The patient, a young adult, was referred from the Institute of Medical Genetics, Wolfson Medical Centre, Holon, Israel in July 2019, where the genetic diagnosis was first made (D.L., M.M., K.Y., M.G.). We previously reported the clinical characteristics of five Yakutian patients (Pavlova, 2019). Retrospective non-identifiable clinical data were analysed according to local and international ethical regulations. Informed consent was obtained from the parents of the patients in accordance with the standards of the Declaration of Helsinki. The study in the present case, patient P6 was approved by the Ethics Committee of The Institute of Medical Genetics, Wolfson Medical Centre, Israel. Parents of the patient consented for publication. The patient P7 was referred for diagnostic advice, the urine sample was obtained with patient consent for clinical investigation.

Detailed information about the samples and biochemical analyses is described in Supplementary information.

### Next Generation Exome and Sanger sequencing

The description of sequencing analysis is provided in Supplementary information.

### Cell culture and immunoblotting

Healthy adult and neonatal fibroblasts were obtained from Sigma (Catalogue numbers 106-05A; 106-05N) and normal adult fibroblasts from Lonza (Catalogue number CC2511). Patients’ and healthy control dermal fibroblasts were cultured in Minimum Essential Medium Eagle (MEM) (Sigma) or Nutrient media F10 supplemented with 10% -20% foetal bovine serum (FBS) (Sigma), 1 mM Sodium Pyruvate (Sigma), 2mM Glutamax-1 (Gibco), 100IU/mL penicillin, 100μg/mL streptomycin (Invitrogene) at 37°C in a humidified atmosphere of 5% CO2.

Patients’ and control fibroblast cell lines were tested and confirmed to be free of mycoplasma species by Eurofins Mycoplasma service.

Methods used for immunoblotting are supplied in Supplementary Information.

### Electron Microscopy

The detailed descriptions for horseradish peroxidase (HRP) uptake to flood the fibroblast endocytic system, fixation, staining and cutting of ultrathin sections are provided in the Supplementary Information. For stereological analysis, 50 images per sample were collected at a magnification of x890 using an unbiased raster pattern (Griffiths, 1993). The volume fraction in individual cell profiles of the cell cytoplasm was determined by point counting using a 10μm^2^ grid overlay and the volume fraction of endocytic organelles containing HRP was determined using a 2μm^2^ grid overlay using NIH Image J (Version 2.0.0-rc-69/1.52i).

### Confocal microscopy

Cells were grown on 4 well glass slides (Millipore) in normal growth medium for 24 hours. After fixation the cells were, permeabilized using the fixation/permeabilization kit (BD Biosciences catalogue 554714) and incubated with primary mouse anti-human LAMP2 (Abcam) or EEA-1 (BD Biosciences) monoclonal antibodies and an Alexa Fluor 488 conjugated goat anti-mouse IgG (H+L) antibody. The slides were mounted using ProLong Gold antifade Mountant with DAPI nuclear stain (Thermo Fisher Scientific). The slides were examined using a Leica Sp5 confocal microscope and serial 0.3-0.5μm Z-stacks images per field (5-7 cells) were acquired. To analyse acidic lysosomes *in situ*, the cells were incubated with LysoTracker red DND-99 (Thermo Fisher Scientific) and examined by confocal imaging. Images were analysed using ImageJ software.

### Lactosyl ceramide trafficking

Cells were incubated with 5μM BSA-BODIPY-C_5_lactosylceramide(LacCer) as previously described (Pavlova, 2019) (Supplementary Information methods).

## RESULTS

### Clinical and biochemical characteristics of an adult patient with mucopolysaccharidosis plus disease

The patient [designated Patient P6], a young adult of Southern European/Mediterranean origin has been clinically monitored since the age of 2-3 years when psycho-motor developmental delay was noted. Abdominal examination revealed hepatosplenomegaly and proteinuria in early childhood. The disease progressed slowly: mucopolysaccharidosis like features, skeletal deformities developed by the age of 6-10 years, (figure 1, A-E; table 1). The patient had a moderate intellectual disability that reflect autism spectrum syndrome. Semi-quantitative analysis of urinary glycosaminoglycans by two-dimensional electrophoresis on a specimen obtained in adulthood, showed increased heparan and dermatan sulphate as these had previously been recorded in patients, designated Patients P1 and P2, harbouring the R498W mutation in the VPS33A protein (Pavlova, 2019) (figure 1, F). In addition, patient P6 had increased urinary hyaluronic acid – a non-sulphated glycosaminoglycan which was not clearly observed in samples of three Yakutian patients with the R498W mutation (Pavlova, 2019). Urine sialylated conjugates and oligosaccharides were not increased. Enzymatic activities of relevant lysosomal hydrolases were normal, thereby, excluding diagnosis of MPS I and II, III A, B, C, D, MPSVI, GM1 and GM2 gangliosidoses, Pompe disease and sialidosis.

**Table 1.**
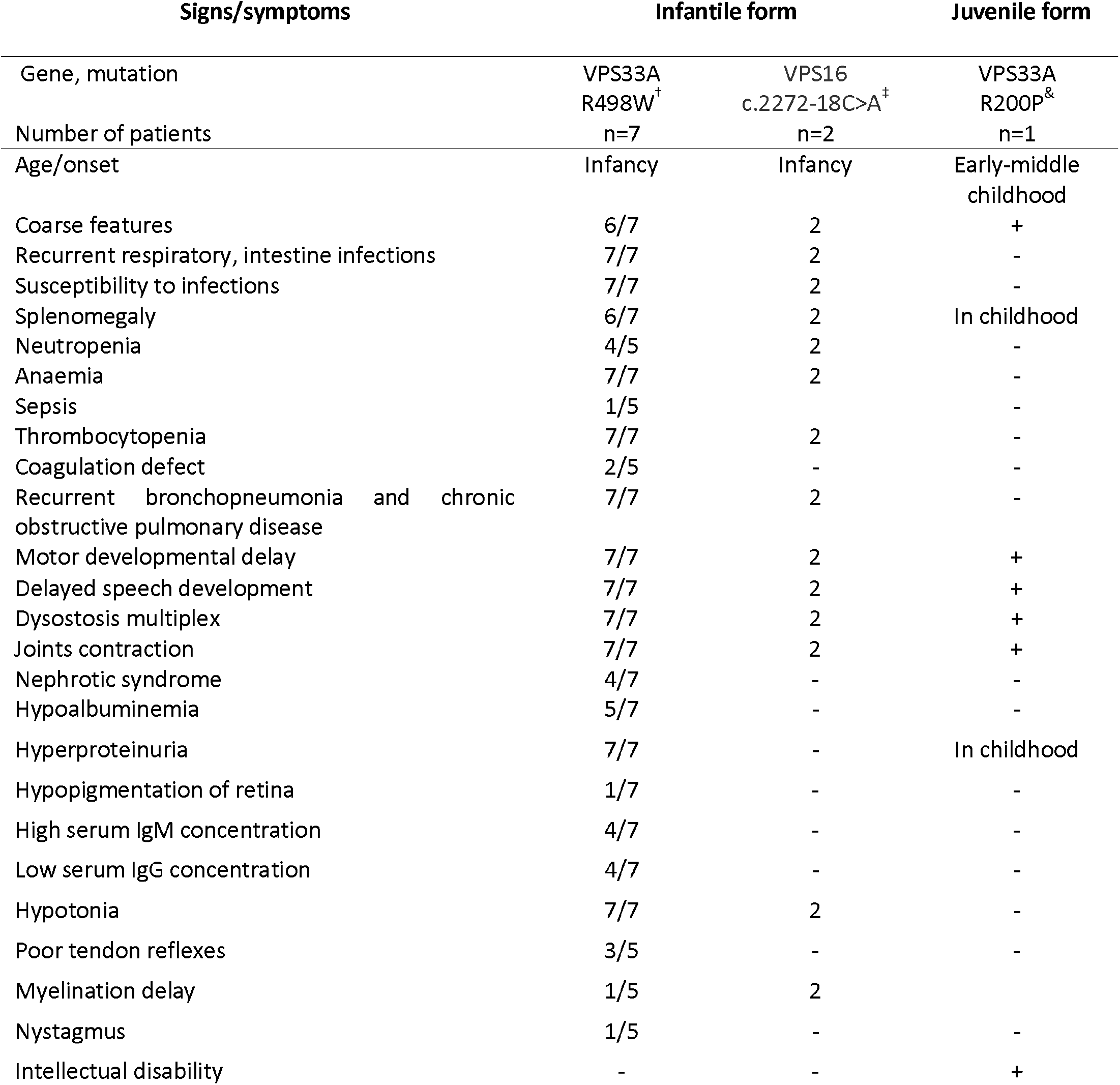
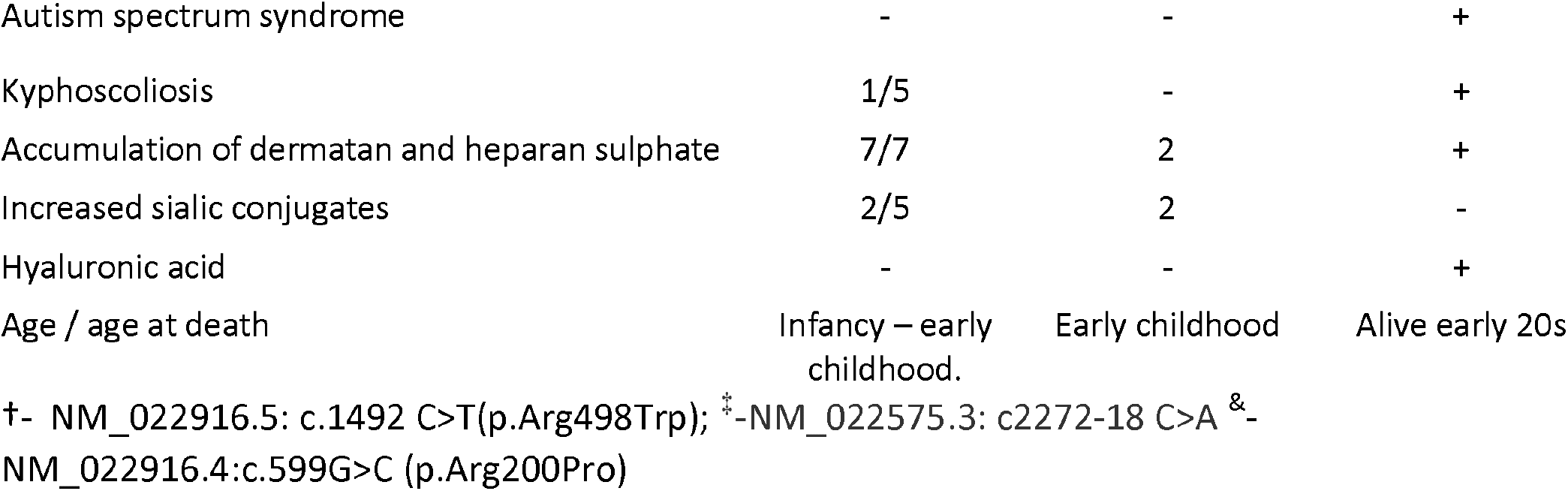
Clinical signs and features in patients with mucopolysaccharidosis plus disease.

**Figure 1.**
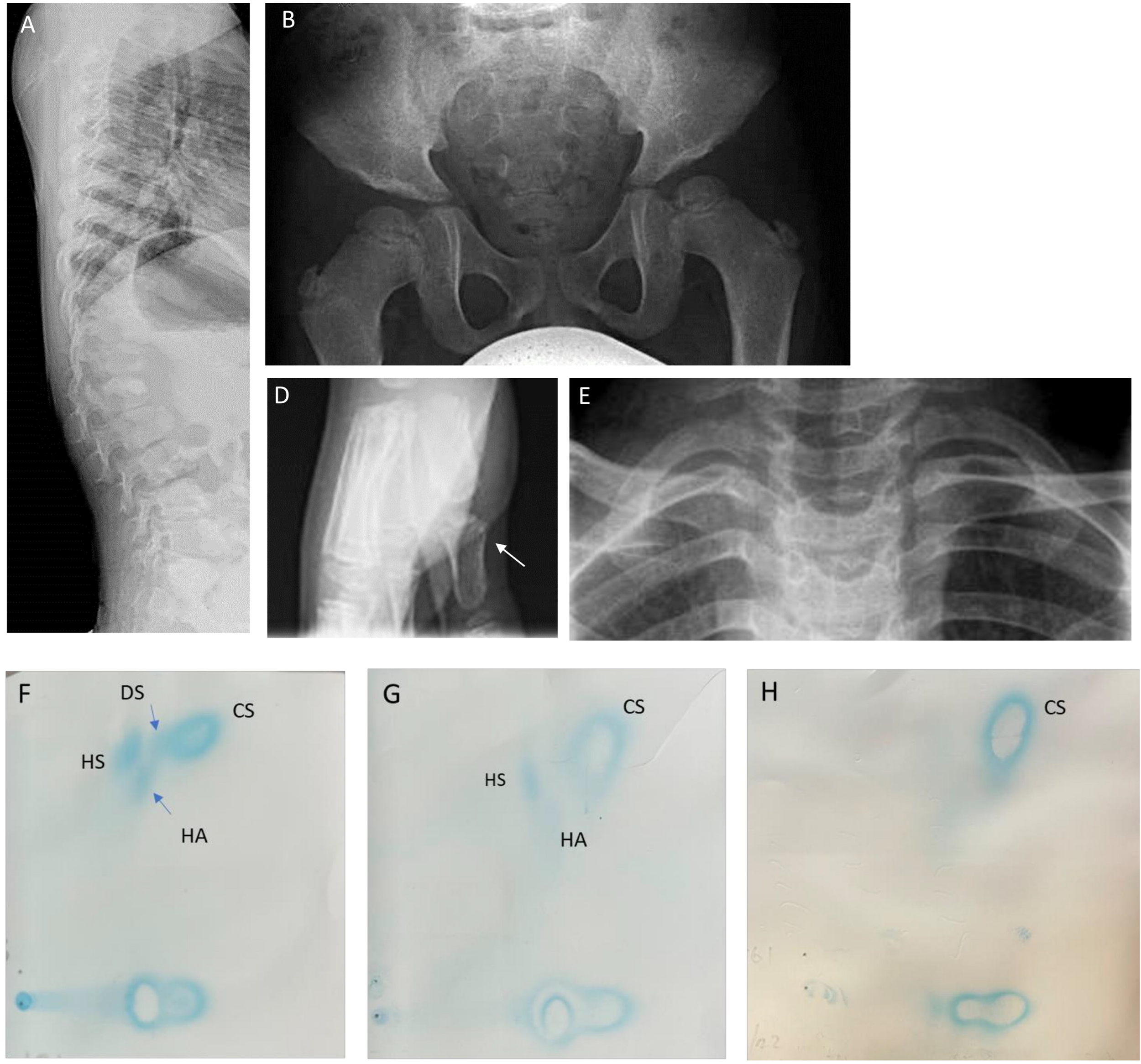
Skeletal complications and two-dimensional electrophoresis of urinary GAGs in patients with a juvenile form of mucopolysaccharidosis plus disease. **A**, Lateral spinal radiograph of patient (P6) in middle childhood showing flattened and oval-shaped vertebrae; **B** Radiograph of pelvic showing flattening acetabula obtained in early childhood; **C**, Patient (P6) in late adolescence showing coarse facial features, ptosis, short neck; **D**, Radiograph of foot in early childhood: widening of metatarsal bones; E, Broad and short claviculae on cervical spine radiograph of patient P6 in late childhood. **F**, Patient P6 urinary GAG pattern showed increased heparan (HS), some dermatan (DS) sulphates and hyaluronic acid (HA). **G**, Patient P7 urinary pattern with some hyaluronic acid (HA). **H**, Healthy individual with normal pattern of GAG.

Another patient of (designated patient P7) was diagnosed with mucopolysaccharidosis plus disease. This patent presented with developmental delay, multiple skeletal deformities, and mitral stenosis. Our urinary GAG analysis did not detect increased heparan, dermatan sulphates, sialylated conjugates and oligosaccharides but showed an increase of hyaluronic acid (figure 1, G).

### Next generation and Sanger sequencing analysis

Next generation exome sequencing of the DNA from patient P6 identified a missense variant NM_022916.4: c.599 G>C in the *VPS33A* gene. This caused replacement of arginine by proline at position 200 of the VPS33A protein (R200P; p.Arg200Pro). Sanger sequencing analysis confirmed that the patient was homozygous and both parents were heterozygous carriers (Supplementary figure S1). In addition, the patient’s healthy siblings were heterozygous carriers indicating autosomal recessive inheritance. Exome sequencing of patient P7’s DNA identified the same missense variant NM_022916.4: c.599 G>C in the VPS33A gene (Krzysztof Szczaluba, Anna Tylki-Szymanska, personal communication).

### Endosomal/lysosomal impairment caused by mutations p.R200P and p.R498W in VPS33A

To investigate intracellular morphological changes, skin fibroblasts from the VPS33A^R200P^ patient P6 were cultured *in vitro*. We observed enlarged vacuoles by electron microscopy of the VPS33A^R200P^ patient-derived fibroblasts as we had previously reported in fibroblasts from VPS33A^R498W^ patients P1 and P2 (Pavlova, 2019). To quantify the enlargement of compartments along the whole endocytic pathway (i.e. all endosomes, endolysosomes and lysosomes), we allowed the cells to endocytose horseradish peroxidase (HRP) continuously for 24h; the volume fraction of HRP-laden endocytic organelles in electron microscopy images of individual cells was determined by a lattice overlay method of stereological sampling (see Methods; figure 2 and Supplementary figure S2). We compared the volume fraction of HRP-positive endocytic organelles in the cytoplasm of the VPS33A^R200P^ fibroblasts (P6, mean 6.2±0.8%) with the volume fraction in fibroblasts from two VPS33A^R498W^ patients (P1, 4.0±0.6%; P2, 3.3±0.6%) as well as neonatal (1.6±0.4%) and adult (2.3±0.3%) control fibroblasts (Figure 2). These data confirmed the enlargement of endocytic organelles in fibroblasts of VPS33A^R498W^ patients, previously inferred from counts of enlarged vacuoles: comparable enlargement was found in the cells from the patient with the VPS33A^R200P^ variant.

**Figure 2.**
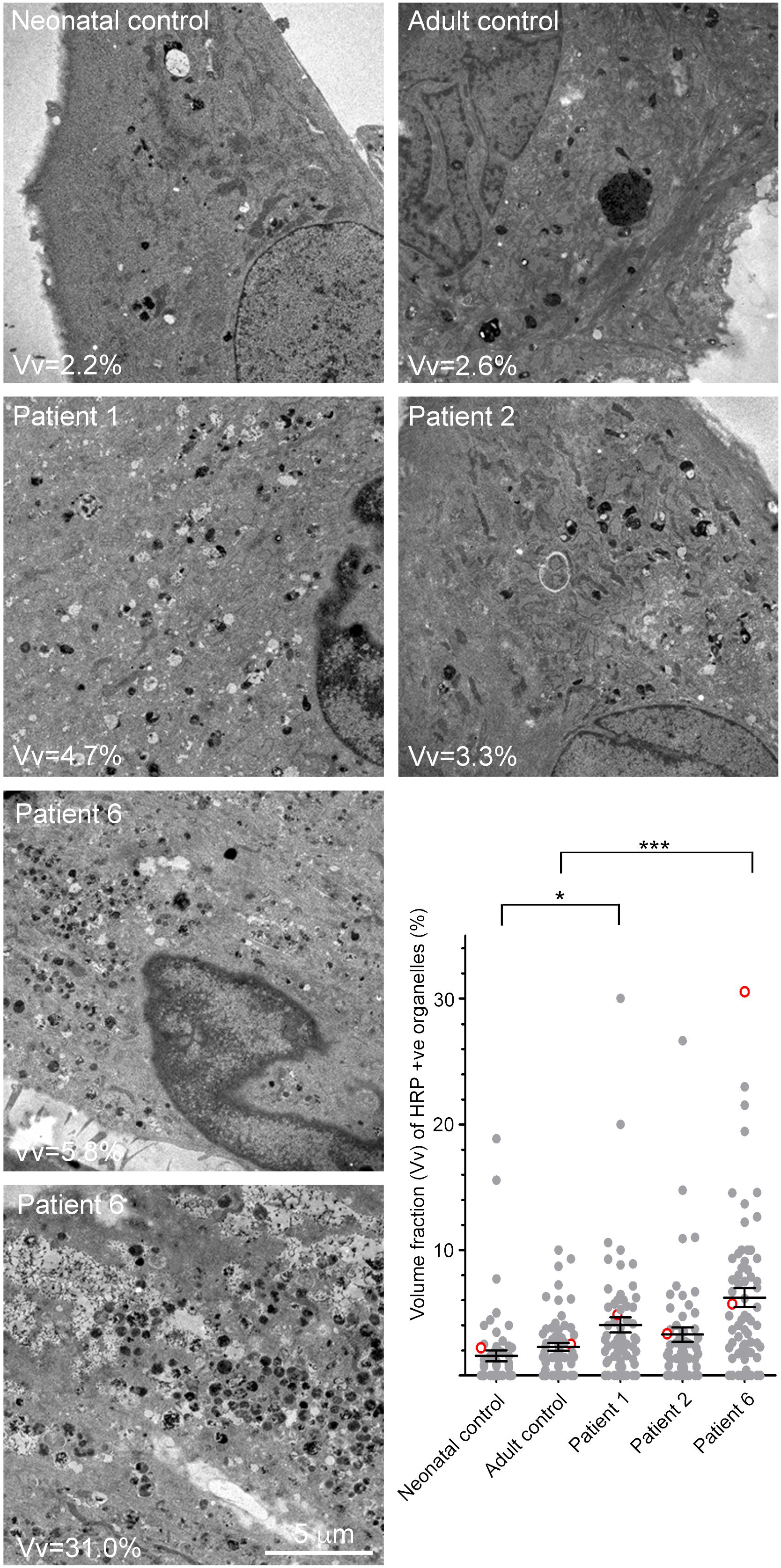
HRP positive endocytic organelles in patient-derived fibroblasts. Electron micrographs of thin sections of control and patient-derived fibroblast cells which had endocytosed medium containing 2mg/ml HRP continuously for 24 h to flood the endocytic system. The volume fraction (Vv) occupied by HRP +ve organelles is shown on each micrograph. The graph shows the volume fraction of HRP +ve organelles in >60 cell profiles in control and patient-derived fibroblasts and the open circles (red border) indicate the position within the data set of the representative electron micrographs shown in this figure.

Further evidence of altered structure and function of the endocytic compartments in VPS33A^R200P^ cells was obtained by fluorescence microscopy. Total immunofluorescence of early endosomal antigen 1 (EEA1) was markedly increased in VPS33A^R200P^ patient-derived fibroblasts (P6, figure 3, A, B) as had previously been observed in VPS33A^R498W^ fibroblasts derived from patients P1 and P2 (Pavlova, 2019). In addition, quantification of EEA1 positive puncta showed a significantly increased proportion of EEA1 positive organelles with an enlarged diameter (1-1.5μm) in the VPS33A^R200P^ patient’s fibroblasts compared with control cells (figure 3, C). Conversely, the proportion of EEA1 positive puncta with a diameter less than 1μm was greater in control fibroblasts (figure 3, C). In contrast to EEA1, but in agreement with previous data from VPS33A^R200P^ patient-derived fibroblasts (Pavlova, 2019), the fluorescence intensity of the lysosomal marker, LAMP2, was not significantly different from that in control fibroblasts (figure 3, A). Incubation of the VPS33A^R200P^ patient-derived fibroblasts with the acidotropic dye LysoTracker Red DND-99, resulted in increased total fluorescent intensity in VPS33A^R200P^ fibroblasts (figure 3, A) compared with control cells, as previously reported for LysoSensor blue dextran DND-167 in VPS33A^R498W^ fibroblasts (Kondo, 2017; Pavlova, 2019). This is consistent with a lower pH of the endosomal/lysosomal compartments in the patient-derived cells. Taken together, the findings on microscopy indicate altered morphology and dysfunction of endosomal/lysosomal compartments in fibroblasts derived from patients with either VPS33A mutation.

**Figure 3.**
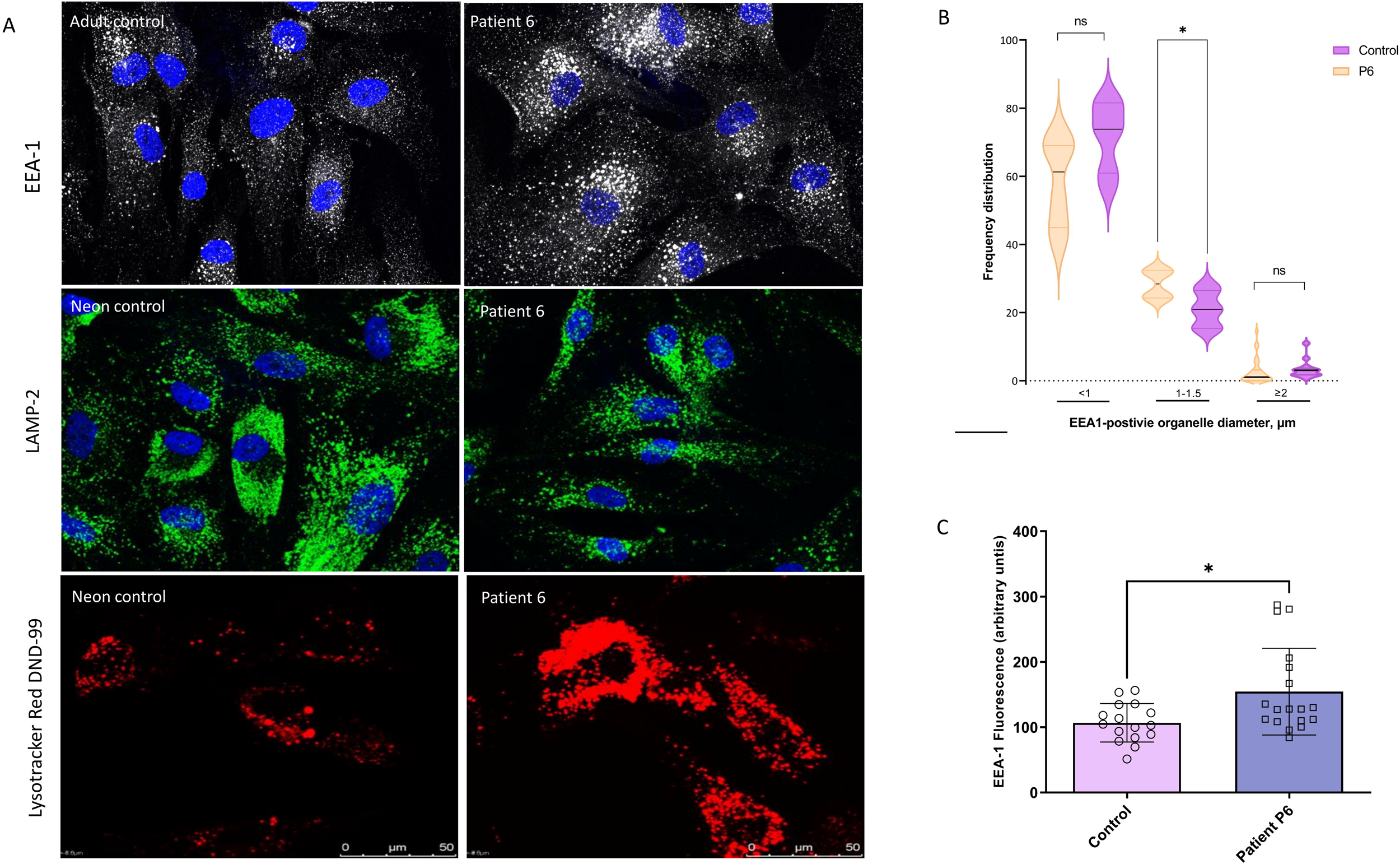
Early endosomal antigen (EEA1) and late lysosomal markers in patient-derived fibroblasts. **A**, Representative confocal images of cultured patient P6 and control fibroblasts stained with early endosomal marker EEA1, lysosome-associated membrane protein LAMP2 antibody and secondary anti-human IgG Alexa-488 (white or green respectively) and DAPI (blue); representative confocal images of patient P6 and neonatal control show increased acidotropic probe LysoTracker red DND-99 fluorescence intensity. **B**, Distribution of EEA1 positive organelles accordingly the puncta diameter bin <1μm, 1-1.5μm and >2μm quantified in six cells of each control and P6 fibroblasts. Violin graphs show the median and quartiles of the quantified EEA1 positive organelles. **C**, Integrated density of EEA1 fluorescence in the patient P6 and control cells. *-p<0.05. The fluorescence intensity per cell was quantified in images of maximum intensity Z-projections. The cellular area, the integrated density and the mean gray values were analysed. Measurements of regions without fluorescence were used for background subtraction. The net average fluorescence intensity per pixel, expressed as corrected total cell fluorescence (CTCF), was calculated for each cell.

### The effect of substitution of arginine 200 by proline on VPS33A protein stability

Examination of the crystal structure of human VPS33A in complex with a fragment of VPS16 (Graham, 2013) showed that the residue arginine 200 is localised in the same domain 2 as the residue arginine 498 (Supplementary figure S3). Arginine 200 is in a helix and forms an intramolecular salt bridge with glutamic acid 525 (Figure 4, A). Replacement of arginine 200 by proline is predicted to disrupt the helix, salt bridge and local ionic network, resulting in destabilisation of the folded protein thereby accelerating proteasomal degradation. Interspecies alignment analysis of the protein sequence showed that the arginine 200 and glutamic acid 525 residues are preserved in vertebrates with evidence of evolutionary conservation from amoebae to vertebrates (figure 4, B).

**Figure 4.**
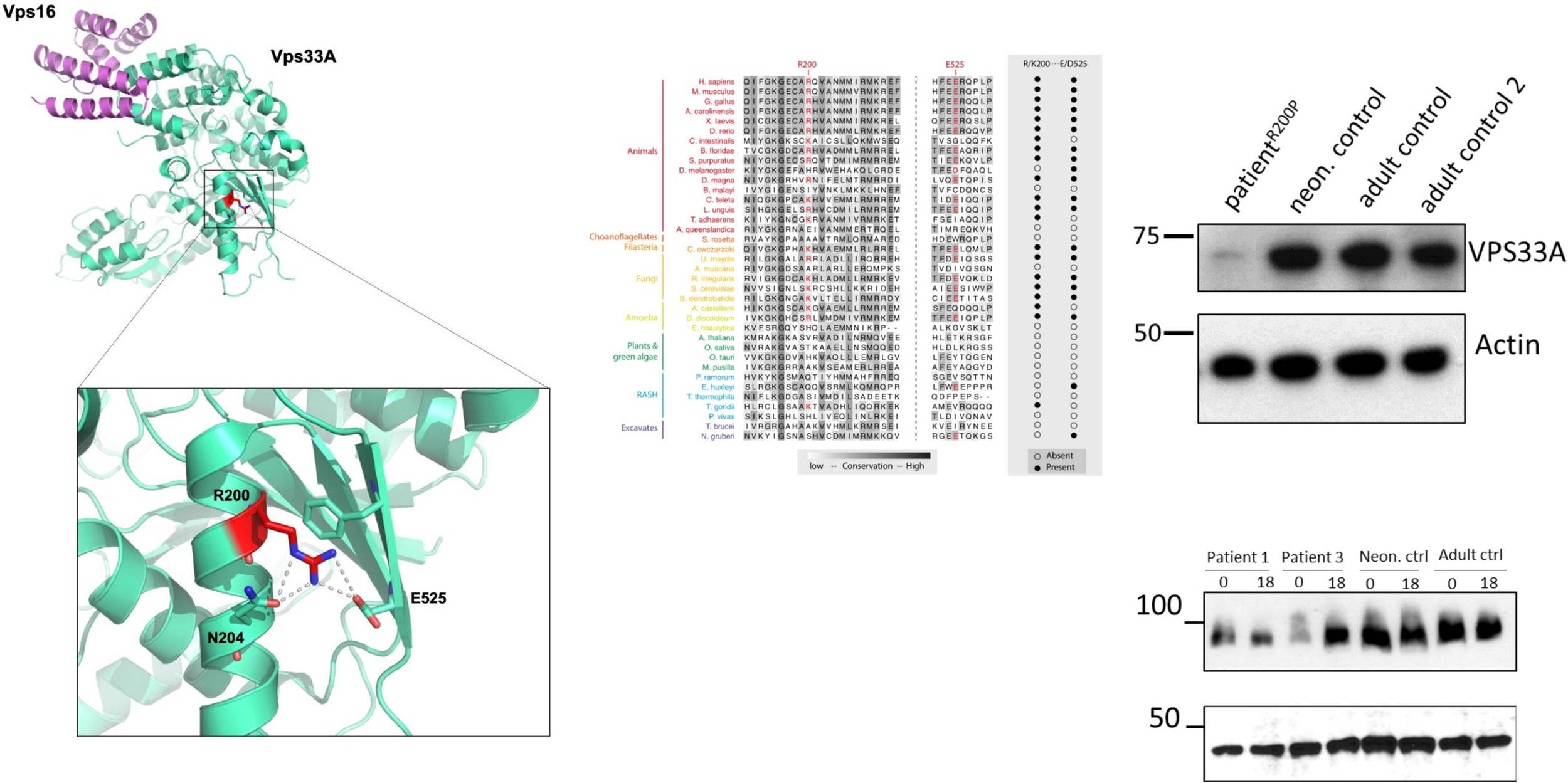
The R200P mutation affects stability of the VPS33A protein. **A**, Cartoon representation of the crystal structure of human VPS33A (green) in complex with VPS16 (Graham, 2013; Wartosch, 2015). Arginine R200 (highlighted red) is located in a helix and forms a salt bridge with glutamic acid 525. **B**. Phylogenetic tree and alignment of VPS33A sequences shows conservation of arginine 200 and glutamic acid 525 residues in vertebrates and amongst species from amoebae to higher vertebrates. **C**. Immunoblot analysis showing reduced abundance of VPS33A protein in patient fibroblasts (P6) compared to neonatal (neon) and two adult control fibroblasts. **D**. In fibroblasts of patient (P1) with R498W and patient (P6) with R200P mutated VPS33A, reduced protein abundance was also observed for the HOPS/CORVET subunit VPS16. Actin was used as a loading control. Fibroblasts of patient P1 and patient P6 were incubated with 100μM bortezomib (PS-341, Stratech, catalogue number S1013) for 18 hours in normal cell culture media (MEM supplemented with 10% (v/v) FCS, (2mM glutamine, 100U/mL penicillin and 100μg/mL streptomycin) and cell lysates analysed by immunoblotting with polyclonal rabbit anti-human VPS16 antibody and secondary HRP-conjugated anti-rabbit IgG or anti-actin antibodies.

Immunoblotting of protein lysates obtained from patient-derived VPS33A^R200P^ fibroblasts revealed a decreased abundance of full-length (65kDa) VPS33A compared with three healthy control fibroblast lysates (figure 4, C). These data are consistent with substitution of residue arginine 200 by proline resulting in reduced protein stability and proteolysis. Destabilization of VPS33A is predicted to impair assembly/stability of the CORVET and HOPS complexes (Wartosch, 2015). In support of this argument, abundance of the CORVET/HOPS component VPS16 as detected by immunoblotting was also reduced in lysates obtained from VPS33A^R498W^ and VPS33A^R200P^ (P1 and P6 respectively) lysates (figure 4, D). Moreover, incubation of patient-derived fibroblasts with a proteasome inhibitor, bortezomib, for 18 hours, increased the steady state concentration of VPS16 in the lysates close to that in control cells (figure 4, D).

### Delayed BODIPY-Lactosylceramide trafficking in VPS33A^R200P^ patient derived fibroblasts

We previously reported a delayed trafficking of endocytosed BODIPY-Lactosylceramide C_5_-lactosylceramide (BODIPY-LacCer) in fibroblasts obtained from patients with VPS33A^R498W^ (Pavlova, 2019). To investigate whether VPS33A^R200P^ has a similar effect on lactosylceramide trafficking we conducted BODIPY C_5_-lactosylceramide (BODIPY-LacCer) pulse-labelling and chase experiments in fibroblasts from patients and control fibroblasts (Pavlova, 2019; te Vruchte, 2004). As shown in VPS33A^R498W^ fibroblasts (Pavlova, 2019), when compared with the signal in control fibroblasts BODIPY fluorescence in the VPS33A^R200P^ patient P6 cells was observed in more peripheral punctate structures, consistent with its accumulation in endocytic organelles (figure 5). As previously reported in fibroblasts of VPS33A^R498W^ patients, quantification of BODIPY-LacCer fluorescence confirmed that there were fewer peripheral puncta in the VPS33A^R200P^ patient’s cells exposed to treatment with bortezomib (100μM, 18 hours) (figure 5, A). This delayed glycosphingolipid trafficking in patient P6 fibroblasts was partially corrected by treatment with the protease inhibitor, in three independent experiments (figure 5, C).

**Figure 5.**
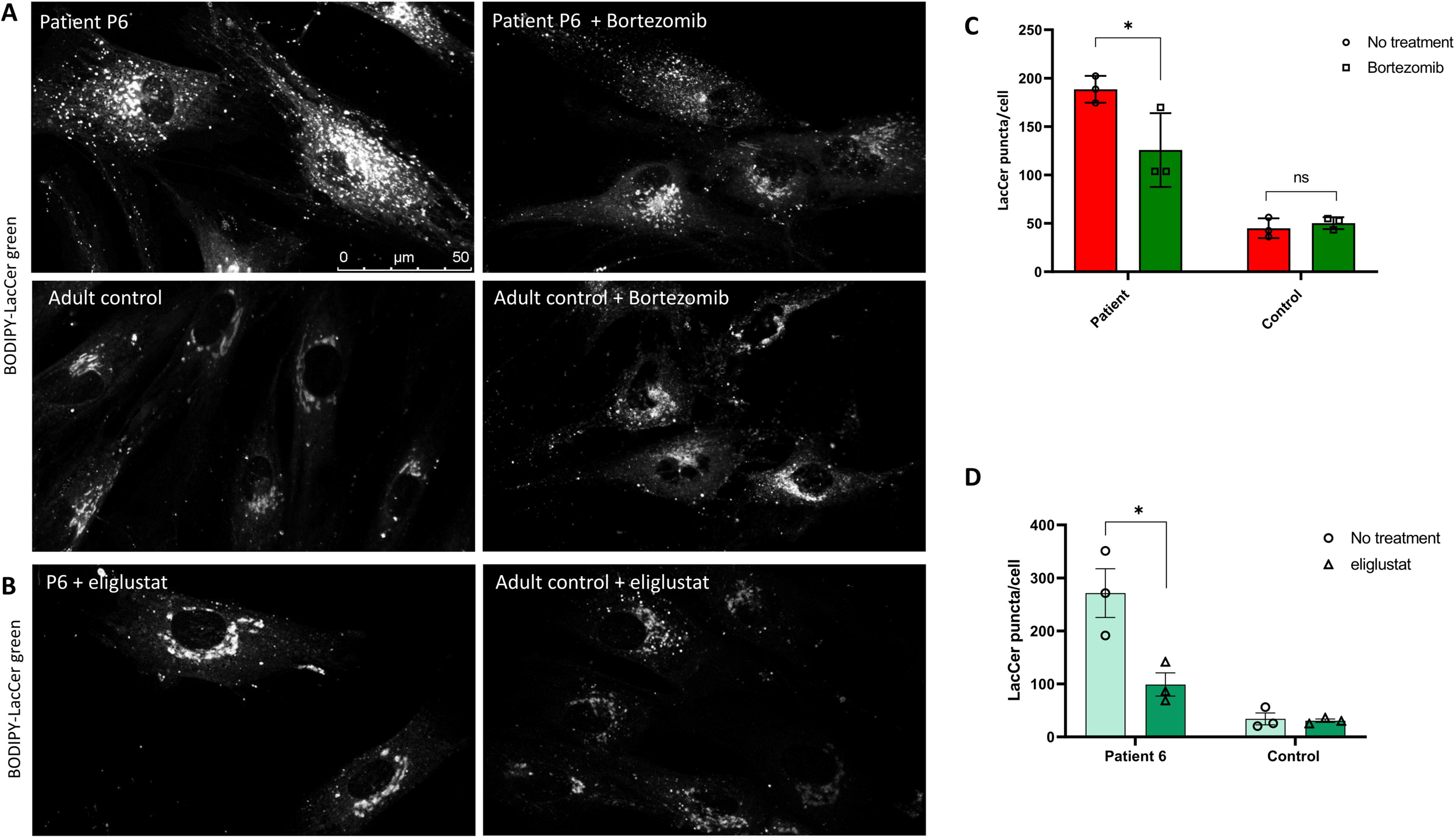
Abnormal BODIPY-LacCer trafficking in patient-derived fibroblasts is corrected by proteasome inhibitor bortezomib and improved by inhibitor of β-glucosylceramide synthesis, eliglustat tartrate. Patient-derived or control fibroblasts were grown on 35-mm glass-bottom tissue culture dishes in normal media supplemented with 100μM of Bortezomib (PS-341, Stratech, catalogue number S1013) for 18 hours. **A**, Fibroblasts were labelled with BODIPY-LacCer and endocytosed BODIPY-LacCer was observed after 3 hours uptake. **B**, Patient-derived (P6) and control fibroblasts were incubated with a specific inhibitor of UDP-glucosylceramide synthase, 50nM eliglustat tartrate for 24 hours. Representative images of patient-derived cells and control cells labelled with BODIPY-LacCer and observed on confocal microscope. **C**, Quantification of BODIPY-LacCer puncta in the patient-derived (P6) and control fibroblasts was performed in ≥10 cells per each line from three independent bortezomib experiments. Data presented as Mean ± SEM, * - p<0.05, NS - not significant. Patient-derived and control fibroblasts were incubated with a specific inhibitor of UDP-glucosylceramide synthase, 50nM Eliglustat tartrate for 24 hours. **D**, Quantification of BODIPY-LacCer puncta in patient P6 and control fibroblasts in ≥10 cells from three independent eliglustat experiments. 5-10 single live cell images (each containing 3-5 cells) were analysed using ImageJ software. For each image, an 8-bit grey scale image was generated using thresholds removing background signals. Pixels clearly corresponding to Golgi fluorescence were selected and removed from the images and puncta containing the remaining pixels were counted. Data presented as Mean ± SEM, * - p<0.05.

### Substrate reduction therapy improves Lactosylceramide trafficking in VPS33A^R200P^ patient-derived cells

To investigate if reduction of glycosphingolipid biosynthesis could improve the endosomal-lysosomal recycling of fluorescent lactosylceramide we treated patient P6 fibroblasts with 50nM eliglustat and miglustat, N-butyldeoxynojirimycin for 24 hours as previously described (Pavlova, 2019) and pulse-chase labelled the fibroblasts with BODIPY-LacCer. Also, as was previously observed with VPS33A^R498W^ patients’ fibroblasts, exposure to 50nM eliglustat and 100μM miglustat, redistributed BODIPY fluorescence principally to the Golgi area (figure 5B and data not shown). This indicates that inhibition of de novo glycosphingolipid biosynthesis reduces intracellular accumulation of glucosylceramide and improves endocytic trafficking in VPS33A^R200P^ fibroblasts (figure 5, B and D).

## DISCUSSION

The recently recognized class of membrane trafficking disorders (Garcí a-Cazorla, 2022) such as that due to VPS33A mutations - mucopolysaccharidosis plus, [caused by deficiency of the HOPS complex], has pleiotropic clinical features. The infantile form manifests in the first year of life with delayed psychomotor development, failure to thrive, diffuse muscle hypotonia, joints contractures, dysostosis multiplex and rapid progression of multi-system disease affecting respiratory, immune, cardiovascular, haematological, gastrointestinal, renal and central nervous systems. Patients suffer life-threatening episodes of sepsis related to immunodeficiency and often require support that includes assisted ventilation, parenteral nutrition and blood transfusions. Patients with the infantile form of the disease died with multi-system failure triggered by infection within the first 6 years of life. Here we describe an attenuated juvenile onset form of mucopolysaccharidosis plus that is due to inheritance of a previously unreported homozygous variant p.R200P in VPS33A protein, a core subunit of the HOPS and CORVET tethering complexes identified in an adolescent patient with long-standing intellectual disability, skeletal deformities and evidence of mild cardiac disease (Dursun,2017; Kondo, 2017; Pavlova et al., 2019).

Of note, two patients have been lately reported with an overlapping phenocopy of mucopolysaccharidosis plus disease caused by genetic deficiency of VPS16, another subunit of the HOPS and CORVET tethering complexes (Sofou, 2021) to which VPS33A is recruited by binding. These patients had progressive psychomotor regression, delayed myelination, and mucopolysaccharidosis-like features apparent in infancy (table 1). Both patients were homozygous for the same intronic variant (NM_022575.3: c.2272-18C>A) in VPS16, that generates a hypomorphic allele causing the deficiency of both subunits VPS16 and VPS33A. These patients also had marked systemic manifestations in common with infants with the R498W mutation in VPS33A and features shared with Chédiak-Higashi syndrome including neutropenia, splenomegaly, thrombocytopenia, vacuolised peripheral lymphocytes and dense azurophilic granules in myeloid progenitor cells of bone marrow with susceptibility to infections. Patient-derived fibroblasts showed defective endosomal trafficking of transferrin and accumulation of autophagosomes (Sofou, 2021). In this connection, the development of a steroid-responsive acute haemophagocytic histiolymphocytosis recently reported in an infant of Moroccan ethnic background with mucopolysaccharidosis plus and the identified R498W mutation in VPS33A, merits attention. The response to intensive corticosteroid therapy is typical of an impaired innate immunity and possible autoinflammation triggered by infection in patients with mucopolysaccharidosis plus disease (Faraguna, 2022).

Inborn errors of abnormal endocytic trafficking caused by deficiency of VPS33A, VPS16 and potentially all other subunits of the HOPS complex are likely to affect biogenesis of lysosome-related organelles functioning at the immune synapse as well as presynaptic biogenesis in neurons (van der Beek, 2019; van der Welle, 2021; Edvardson, 2015; Sanderson, 2021). Failed recruitment and assembly of HOPS complex subunits on lysosomal membranes due to the defective interactions with small GTPase ADP-ribosylation factor-like protein 8B (Arl8b) is associated with impaired fusion of phagosomes with lysosomes and disturbed phagolysosomal maturation (Khatter, 2015; Sasaki, 2013; Barry, 2012; Garg, 2011; Abo, 1982; Cox, 2022). It moreover delays presentation of a synthetic glycolipid, α-galactosylceramide, to CD1d molecules in the lysosome and impedes microbial killing (Luzio, 2014). Immune deficiency caused by impaired development of NK cells and cytotoxic T cells has been documented in lysosomal biogenesis defects and abnormal endosomal-lysosomal trafficking (Ham, 2014; Orange, 2008; Ballabio, 2020). Finally, the severe nephritis with glomerular sclerosis and proteinuria is likely to be related to vacuolation and effacement of podocytes in which active endocytosis is impaired (Chen, 2013; Bechtel, 2013; Sofronova, 2022).

Unlike patients with the infantile form of mucopolysaccharidosis plus disease due to homozygosity for the R498W mutation in VPS33A, even by early adulthood, the patient P6, who is homozygous for the R200P mutation in the same region of the protein, has not hitherto developed full-blown systemic complications such as glomerulonephropathy, broncho-pulmonary complications associated with respiratory distress, susceptibility to infections or spontaneous haemorrhage. Environmental factors, exposure to infections, vulnerability of the immune system in early childhood or genetic background may explain incomplete clinical penetrance in this monogenic disease and hence a mild clinical phenotype without the severe sporadic infections in genetically predisposed children (Abel, 2010).

At the biochemical level, however, urinary excretion of heparan and dermatan sulphate was raised in our patient, P6 in his 20s - as in patients with neonatal onset disease. These glycosaminoglycans were not, however, detected in the urine sample obtained from younger patient P7, also with the R200P mutation. Of special note, there was also an increased excretion of the non-sulphated glycosaminoglycan, hyaluronic acid, in both patients homozygous for the R200P mutation (figure 1F, G), and this abnormality may occur in infants with the R498W mutation. Hyaluronan receptors (stabilin-1 and stabilin-2) have been identified in electron-dense vesicles in liver sinusoidal endothelial cells and co-localise with EEA1-positive endosomes, suggesting that these receptors mediate clearance of endocytosed hyaluronic acid and possibly other glycosaminoglycans species - dermatan and heparan sulphates (Hansen, 2005). This observation suggests that accumulation of glycosaminoglycans is unrelated to the primary digestive function of lysosomal hydrolases but could result from abnormal intracellular trafficking via the endocytic pathway.

From the practical aspect, it is of paramount importance in the diagnosis of suspected lysosomal diseases with features of mucopolysaccharidosis, that advanced and sensitive methods for glycosaminoglycan analysis are used and that genetic studies should include *VPS33A* in relevant diagnostic panels. The clinical signs of intellectual disability with autism spectrum syndrome merit further studies to understand a role of the HOPS/CORVET complexes in impairment of cognitive skills development and mental retardation. The development of these disabilities in our patient immediately illustrates the need for diagnostic awareness combined with planning for care and clinical monitoring support over childhood and well into adult life. The occurrence of an attenuated VPS33A defect reported here for the first time in a young adult with late onset mucopolysaccharidosis plus disease and slowly progressive course limited to the skeletal system with psychomotor delay, clearly indicates a better prognosis for survival.

Fibroblasts obtained from patients with both forms of mutated VPS33A (R498W and R200P) show vacuolation and enlargement of endosomal and lysosomal compartments along the whole endocytic pathway. The diseased fibroblasts, moreover, showed enhanced fluorescence of Lysotracker dextrans, implying increased acidification and increased fluorescence of EEA1. Increased lysosomal acidification was attributed either to direct regulation of acidification (Kondo, 2017) or as a consequence of an altered balance of the fission and fusion events that can regulate the size and acidity of endosomal/lysosomal compartments (Pavlova, 2019; Bright, 1997). Just as had previously been noted in fibroblasts from VPS33A^R498W^ patients (Pavlova, 2019) we identified delayed trafficking of endocytosed lactosylceramide and its accumulation in endocytic organelles in the VPS33A^R200P^ patient. While this confirms impaired membrane trafficking, a feature observed in several sphingolipid diseases, regardless of their primary defect in lysosomal enzymes, it relates to the effects on endosomal/lysosomal compartments in lysosomal storage diseases and reflects the generally impaired function of the endosomal-lysosomal compartment (Pagano, 2003; Puri et al., 1999; te Vruchte et al., 2004; Pryor et al., 2006; Marks et al., 2013; Bright, 1997). Treatment with bortezomib significantly ameliorates delayed trafficking of fluorescent glycosphingolipid to the Golgi complex (figure 5), providing evidence that an increased steady state concentration of the mutated VPS33A and hence, the assembled HOPS complex, improved intracellular endocytic membrane trafficking.

A recent study using a combination of light and electron microscopy techniques on ultrathin cryosections and systematic analysis of fluorescent organelles to compare with endolysosomal morphology demonstrated that EEA1, usually regarded as an early endosomal marker was also present on a population of later endosomes after loss of Rab5, but where PI(3)P was still present (van der Beek, 2022). Thus, swollen EEA1-positive organelles and increased LysoTracker fluorescence in the R200P and R498W mutant cells are likely to be related to the endosomes and lysosomes where the principal function of HOPS complex has been deficient. However, as previously shown in R498W fibroblasts (Kondo, 2017; Pavlova, 2019), there would be sufficient residual HOPS complex to bring about fusion of catalytically active lysosomes or autophagosomes in which uptake and delivery of fluorescent dextran to endosomes and lysosomes with a complement of active cathepsin B was unimpaired (Pavlova, 2019).

Our findings of accumulation of hyaluronic acid, dermatan and heparan sulphates and increased EEA1 positive endosomes in patients with deficiency of HOPS and potentially CORVET complexes are intriguing and prompt further studies on the dynamic molecular interactions of particular glycosaminoglycans during endocytosis. Here we show that the two mutations described in VPS33A reduce the abundance of the protein and are also associated with diminished amounts of VPS16, another subunit common to the HOPS and CORVET complexes. This finding indicates that the complement of fully assembled HOPS complexes is decreased and that the interaction between VPS33A and VPS16 is necessary for the fusion of endosome and lysosome and autophagosome in living cells as was found in knockout experiments previously conducted *in vitro* (Sofou, 2021).

The young man reported here has a relatively mild phenotype that might be accessible to empirical and ethically approvable therapeutic exploration with known repurposed drugs. It is noteworthy that substrate reduction therapy with eliglustat and miglustat (inhibitors of UGCG, which catalyze the biosynthesis of β-D-glucosylceramide, the precursor of gangliosides and cognate glycosphingolipids), improved glycosphingolipid trafficking in VPS33A^R200P^ fibroblasts (figure 5). These findings immediately suggest that partial correction of the delayed turnover and accumulation of the glycosphingolipids in the lysosomal compartment can be achieved by inhibiting the *de novo* synthesis of gluco-series sphingolipids (Cox, 2017; Pavlova, 2015). Increased glucosylceramide concentration may activate an alternative pathway that converts glucosylceramide to glycosylated cholesterol through transglycosylation catalysed by intracellular GBA2 located outside of the lysosomal membrane (Marques, 2016; Boer, 2021; Ghisaidoobe, 2014). However, it remains unclear as to whether glycosylated cholesterol is formed in cells with VPS33A deficiency as a consequence of defective membrane sphingolipid trafficking.

In summary, we report a juvenile form of mucopolysaccharidosis plus disease caused by a hitherto unknown mutation R200P that affects a conserved residue in the VPS33A protein with an attenuated phenotype and neuropsychological complications manifested by late onset and slowly progressive natural course of this disease. Our study shows that the disease caused by deficiency of VPS33A and destabilisation of HOPS [and potentially CORVET complexes] is characterised by an expanded endocytic compartment and an increased proportion of acidic, swollen endolysosomes. Given the accumulation of glycosaminoglycans and glycosphingolipids due to defective endocytic traffic, we further showed that the approved proteasomal inhibitor, bortezomib, and inhibition of glycosphingolipid synthesis with the approved drugs, eliglustat or miglustat are plausible therapeutic options.

## Data Availability

All data relevant to the study are included in the article or uploaded as supplementary information. Data are available upon reasonable request.

## ACKNOWLEDGEMENTS

We thank the patients and their families for their contribution in this study. We are grateful to Cambridge University Hospitals NHS Foundation Trust and the NIHR Cambridge Biomedical Research Centre for technical support with primary fibroblasts culture and facilities at the Cambridge National Institute for Health Research BRC Cell Phenotyping Hub.

We thank Dr Krzysztof Szczaluba and Professor Anna Tylki-Szymańska for sharing information on the identified missense mutation R200P in patient P7 and sending the urine sample for GAG electrophoresis analysis. We would like to thank the Genome Aggregation Database, dbSNP, 1000 Genomes Project and the groups that provided exome and genome variant data to this resource for comparison.

## CONTRIBUTORSHIP STATEMENT

E.V.P., J.P.L., N.A.B. and T.M.C. conceived the experimental study, D.L., M.M. clinically monitored the patient (P6) and collected the clinical information, K.V. and M.H.G. performed and analysed exome and Sanger sequencing and described the methodology, E.V.P. analysed clinical information, organised the study, carried out cell culture experiments and analysed the results, wrote the first draft of the manuscript, H.J.C., K.L.T. carried out urinary glycosaminoglycans electrophoresis, N.A.B. performed electron microscopy experiments and analysed the data, P.T.M. responsible for phylogenetic analysis and sequence alignments, V.K.D. responsible for the analysis of the structure of VPS33A. E.V.P., J.P.L. and T.M.C. edited the manuscript and all authors agreed to submit the final version.

