## Supplemental Information for "Juvenile Mucopolysaccharidosis plus disease caused by a missense mutation in *VPS33A*"

SUPPLEMENTARY INFORMATION

SUPPLEMENTARY METHODS

### Patients and samples

EDTA blood and urine samples were collected for diagnostic investigations and were sent to the certified diagnostic laboratory for GAG analysis. Fibroblasts were established by primary culture of skin samples obtained from skin biopsy of the patient P6. DNA sample of patient P6 was purified from EDTA blood sample using QiAmp DNA Blood Maxi kit (Qiagen, Netherlands). Total RNA was purified from 1 $x10^6$ patient-derived and adult control fibroblasts using the RNeasy Mini kit (Qi agen). cDNA was synthesized by reverse transcription of 200 ng of RNA using high capacity cDNA Reverse Transcription kit (Appliedbiosystems kit 4368814) Thermofisher).

### Next Generation Exome and Sanger sequencing

Whole exome sequencing was carried out on the patient's DNA. The sample was enriched with Sureselect Human All Exome v.5 kit 50Mb (Agilent, Santa Clara, CA, USA). Sequencing was carried out on HiSeq4000 (Illumina, San diego, CA, USA) as 100-bp paired–end runs. Reads were aligned with the human reference genome (assembly GRCh37/hg19). Pipeline was performed using the Genoox platform based on BWA (version 0.7.16) for read alignment and GATK HaplotypeCaller (version 3.7) and FreeBayes (version 1.1.0) for variant calling.

Dataset files including the annotated information were analysed with the following filtering steps: variants which were called less than 9 times and synonymous variants were removed. Variants were filtered based on allele frequency less than 0.01 according to online databases; dbSNP, 1000G, ExAC and gnomAD. Likely pathogenicity was explored further if the variant was truncating (splicing or non-sense), missense or an in-frame indel. Missense and in-frame indels were considered if they were predicted to be pathogenic by online prediction tools, PolyPhen-2, SIFT and Mutation Taster. Conformation and familial segregation were performed using direct Sanger sequencing (3500 Genetic Analyzer Applied Biosystems).

### Urine mucopolysaccharide analysis

Urinary glycosaminoglycan concentration was determined using the standard dimethylmethylene blue dye method,[1]. Two-dimensional electrophoresis was used to differentiate sulphated and non-sulphated glycosaminoglycan patterns,[2, 3].

**Immunoblotting**

Protein lysates of the patients P1 and P3 and control fibroblasts were extracted in NP-40 buffer (Life Technologies) supplemented with protease inhibitors cocktail (cOmplete$^{TM}$ Mini, Roche Diagnostics). 10-30 µg of protein lysates were heated at 100°C for 5 minutes in Laemmli Sample Buffer (BioRad) supplemented with reducing agent, 2-mercaptoethanol, and loaded in pre-casted 10% Tris-glycine gels (NuSep). SDS PAGE was run in Tris-Glycine SDS buffer. Proteins were transferred to 0.45µm PVDF membranes (Millipore), the membranes were blocked in 5% skimmed milk in TBS-Tween 20 for 1-2 hours, and stained with polyclonal mouse anti-human VPS33A antibodies (Abcam, ab88254) or polyclonal rabbit anti-human VPS16 antibodies (Proteintech) overnight at 4°C, following incubation with horseradish - peroxidase conjugated anti-mouse (Abcam) and anti-rabbit antibodies. Blots were developed with Amersham Prime ECL western blotting analysis system (Cytiva Life Science) accordingly the manufacturer instructions. Membranes were stained with monoclonal anti-actin (A2066; Sigma-Aldrich) antibodies and detected with HRP conjugated anti-mouse antibodies.

**Electron Microscopy**

Cells were grown in 6 well plates and incubated with 2mg/ml horseradish peroxidase (HRP, Type VI; Sigma-Aldrich) for 24 h to flood the endocytic system. They were then washed with PBS and fixed with 2% PFA / 2.5% GA in 0.1M Na Cacodylate buffer pH 7.2 and washed with PBS. The 3,3′-diaminobenzidine (DAB; Sigma-Aldrich) reaction was performed by washing the cells with 0.1M Tris buffer, pH 7.4, and incubating with 1mg/ml DAB containing 0.3% $H_2O_2$ in Tris Buffer, pH 7.4, for 10 min at room temperature in the dark. The cells were subsequently washed with 0.1M Na cacodylate buffer, pH 7.2, post-fixed in 1% osmium tetroxide in 0.1M Na cacodylate buffer, pH 7.2, for 1 h and washed with $H_2O$. The cells were *en bloc* stained with 4% uranyl acetate in $H_2O$ for 1 h, dehydrated through a graded ethanol series and infiltrated first with 50:50 ethanol: Agar 100 resin and subsequently with pure Agar 100 resin (Agar Scientific, Stansted, UK) prior to polymerization of the resin overnight at 60$^{o}$C.

Ultrathin sections of the cells were cut using a diamond knife mounted on a Reichert Ultracut UCT ultramicrotome (Leica, Milton Keynes, UK), collected on EM grids and stained with uranyl acetate and Reynold's lead citrate. The sections were observed in a Tecnai G2 Spirit BIOTWIN transmission electron microscope (FEI, Eindhoven, The Netherlands) at an operating voltage of 60 kV.

**Lactosylceramide trafficking**

5-10 single live cell images (each containing 3-5 cells) were analysed using ImageJ software. For each image, an 8-bit grey scale image was generated using thresholds removing background signals. Pixels clearly corresponding to Golgi fluorescence were selected and removed from the images and puncta containing the remaining pixels were counted. Cells were incubated with proteasome inhibitors (Bortezomib, Stratech), or the substrate reduction agents eliglustat tartrate or miglustat as previously described,[3].

**SUPPLEMENTARY RESULTS**

**Figure S1**: Sanger sequencing of the c.599 G>C; R200P variant in the *VPS33A* (NM_022916.4) gene. Carrier heterozygosity in the parents and homozygosity in the patient were confirmed.

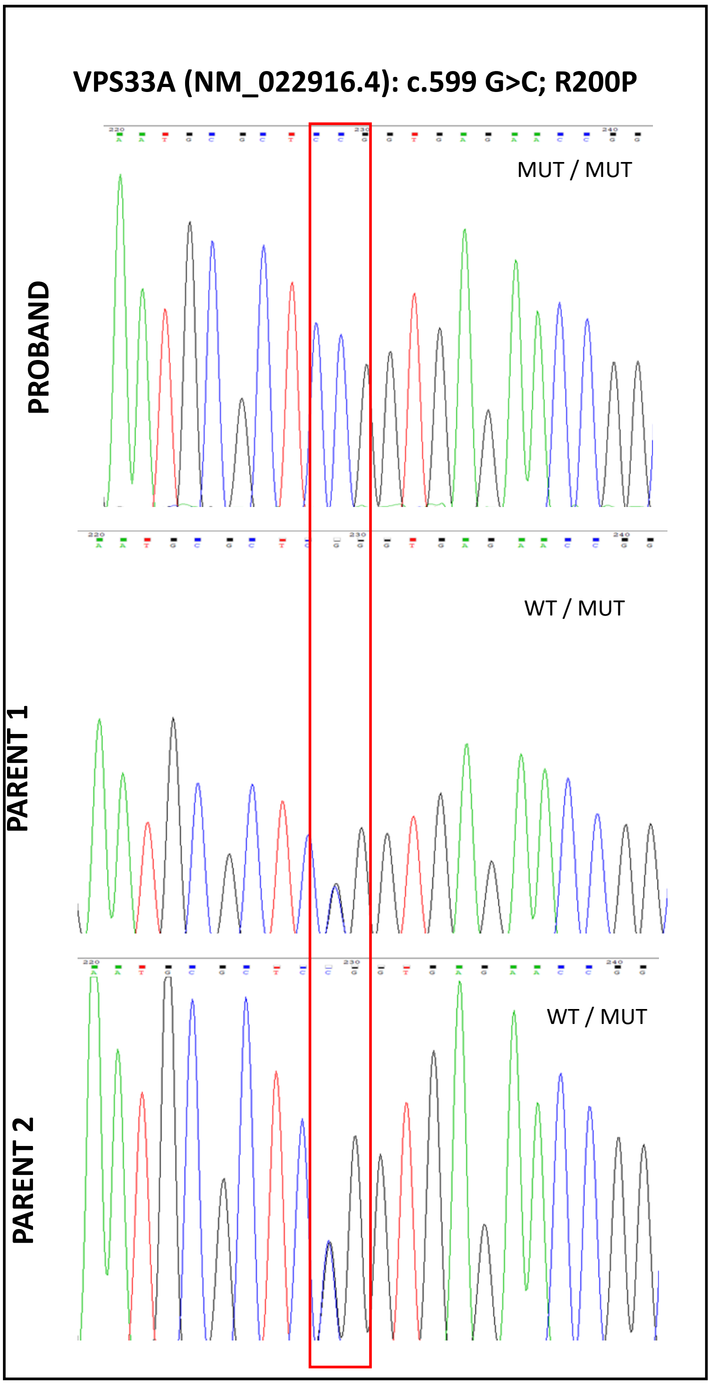

**Figure S2.** Representative micrographs showing stereological sampling with a 2µm2 lattice overlay indicating the volume occupied by HRP laden organelles after 24 h continuous uptake (11 HRP +ve organelle 'hits' underlying lattice points (red); total volume = 22 µm2) and a 10 µm2 lattice overlay indicating cytoplasmic volume (25 'hits' underlying lattice points (cyan); total volume = 250 µm2). Thus, in this example, the volume fraction of cytoplasm occupied by HRP +ve organelles is 8.8%.

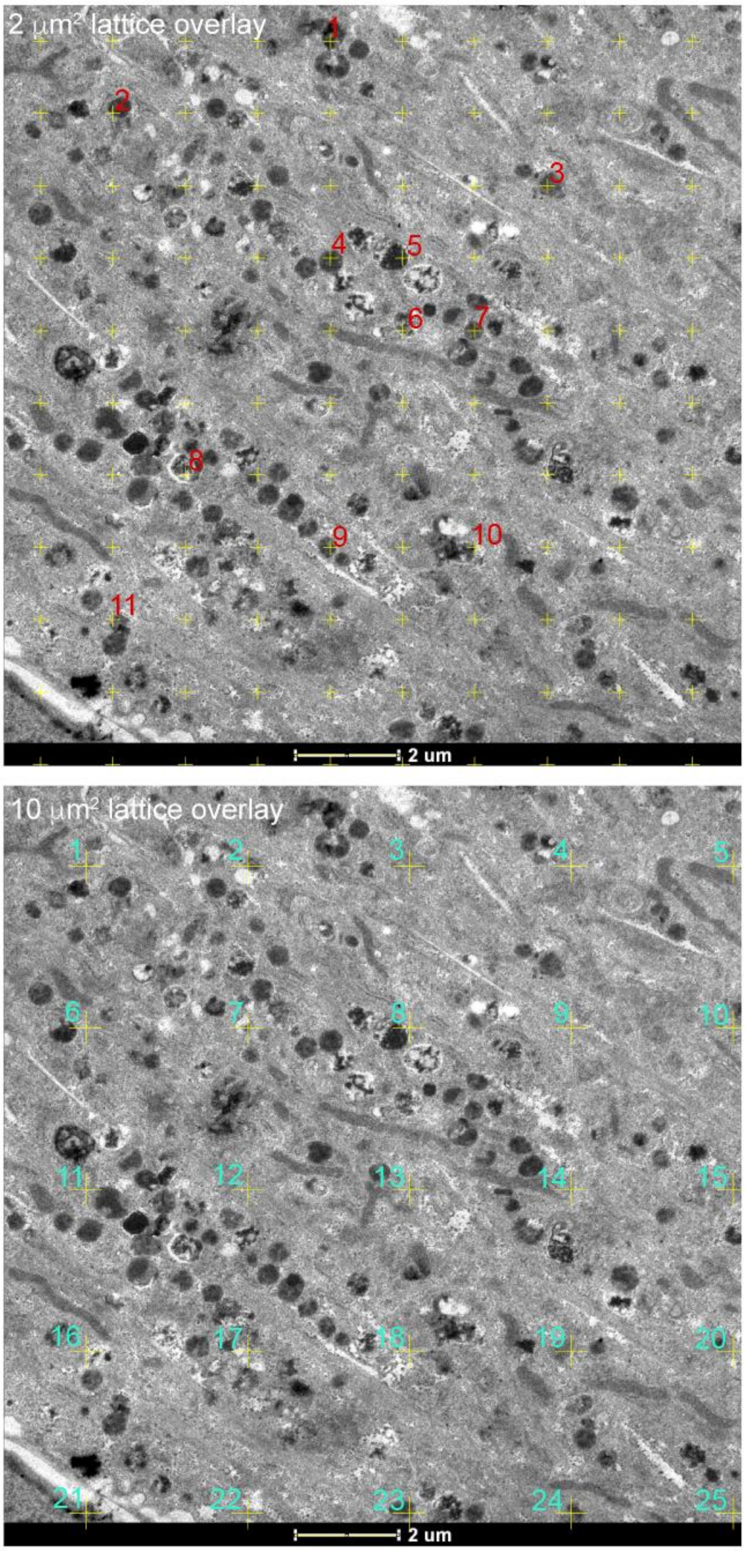

**Figure S3.** Cartoon representation of the crystal structure of human VPS33 A (green) in complex with VPS16. Mutated residues Arginine 200 and Arginine 498 (shown in red) are localised in the same domain 2 of the VPS33A.

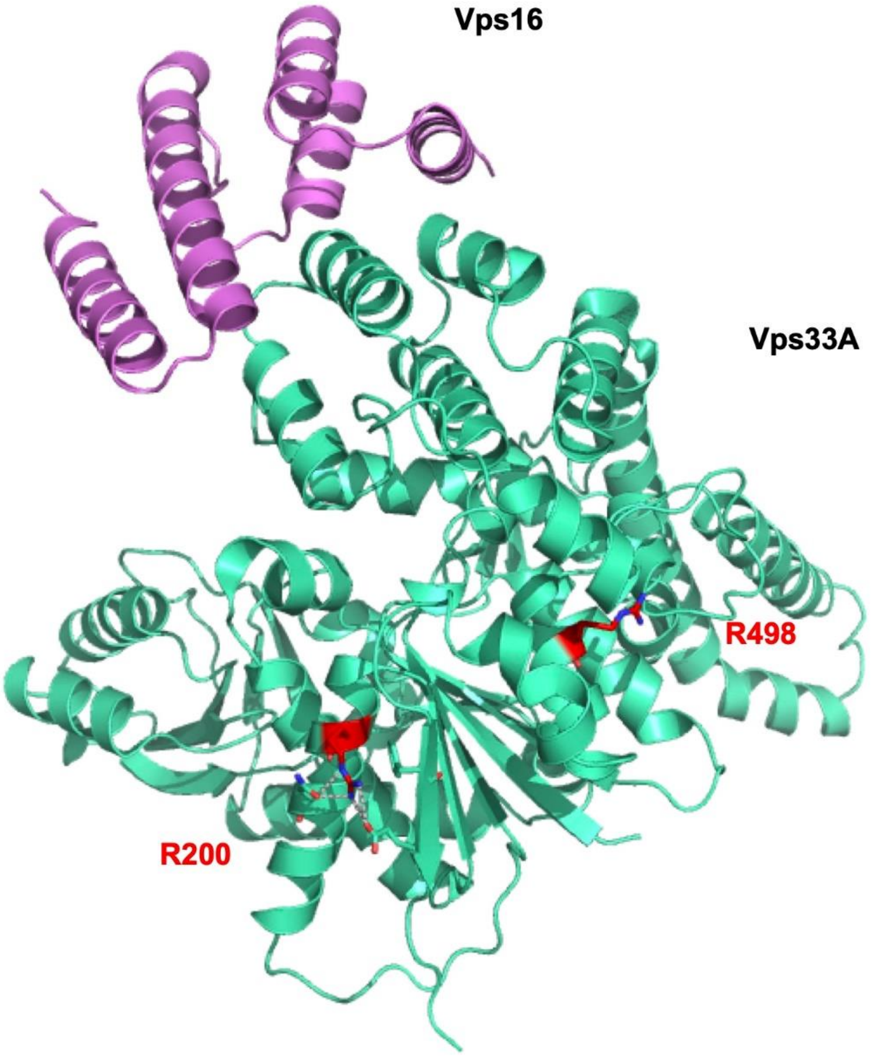
